# Prevalence of missing data in the National Cancer Database and association with overall survival

**DOI:** 10.1101/2020.10.30.20220855

**Authors:** Daniel X. Yang, Rohan Khera, Joseph A. Miccio, Vikram Jairam, Enoch Chang, James B. Yu, Henry S. Park, Harlan M. Krumholz, Sanjay Aneja

## Abstract

**Importance:** Cancer registries are important real-world data (RWD) sources that rely on data abstraction from the medical record, however, patients with unknown or missing data are under-represented in studies that use such data sources.

**Objective:** To determine the prevalence of missing data and its associated overall survival among cancer patients

**Design, Setting, and Participants:** In this retrospective cohort study, all variables within the National Cancer Database (NCDB) were reviewed for missing or unknown values for the three most common cancers in the United States diagnosed from 2006 to 2015. Prevalence of patient records with missing data and their associated overall survival were determined. Data analysis was performed from February to August 2020.

**Exposures:** Any missing data field within a patient record among 63 variables of interest, from over 130 variables total in the NCDB.

**Main Outcome and Measure:** Prevalence of cancer patient records with missing data and associated two-year overall survival

**Results:** A total of 1,198,749 non-small cell lung cancer (NSCLC) patients (mean [SD] age, 68.5 [10.9] years; 569,938 [47.5%] women), 2,120,775 breast cancer patients (mean [SD] age, 61.0 [13.3] years; 2,101,758 [99.1%] women), and 1,158,635 prostate cancer patients (mean [SD] age, 65.2 [9.0] years; 0 [0%] women) were included for analysis. For NSCLC, there were 851,295 (71.0%) patients with missing data in variables of interest; 2-year overall survival was 33.2% for patients with missing data and 51.6% for patients with complete data (p<0.001). For breast cancer, there were 1,161,096 (54.7%) patients with missing data; 2-year overall survival was 93.2% for patients with missing data and 93.9% for patients with complete data (p<0.001). For prostate cancer, there were 460,167 (39.7%) patients with missing data; 2-year overall survival was 91.0% for patients with missing data and 95.6% for patients with complete data (p<0.001).

**Conclusions and Relevance:** Within a large cancer registry-based RWD source, missing data that was unable to be ascertained from the medical record was highly prevalent. Missing data among cancer patients was associated with heterogeneous differences in overall survival. Improving documentation and data quality are needed to best leverage RWD for clinical advancements.

## Introduction

Real-world evidence derived from real-world data (RWD) holds substantial promise to accelerate innovation within oncology. RWD, which includes data on patient health status and/or the delivery of health care collected routinely,^1^ is becoming increasingly relevant due to the high cost and slow pace of randomized clinical trials as well as the growth of near real-time access to electronic health records (EHR) and other digital sources of comprehensive health-related data. RWD sources may represent a flexible, cost-effective way to investigate clinical interventions and can supplement clinical trials. Within oncology, there have been investments in developing RWD sources for clinical evidence generation both at the national level and within professional societies.^2-5^

Cancer registries have long been established as important sources of RWD within oncology for generating insights spanning cancer epidemiology and comparative effectiveness of therapies.^2,6^ Data quality including the completeness of data elements is a major consideration when working with these registries to generate clinical insights. This is particularly germane given emerging evidence suggest that treatment-associated survival outcomes using registry and similar randomized controlled trials are not concordant.^7-9^ There is a critical need to assess the quality of clinical evidence generated from registry and other RWD sources, as well as their adherence to best data practices. Of note, cancer registries rely on trained tumor registrars to abstract and record data from the patient medical record. Lack of quality documentation within the medical record can lead to incompletely abstracted data elements, and therefore lead to unknown or missing data values within cancer registries.^10-12^

While there are a variety of methods to account for missing data, patients with unknown values are likely under-represented in RWD studies, as it is common practice to exclude patients without complete information in variables used for cohort construction.^13-16^ However, because missing data within registries is a surrogate for poor quality documentation, such data may not be “missing completely at random”, and the exclusion of such patients may introduce significant bias. In addition, missing data is also relevant to clinical care as it may reflect important missing clinical information, such as cancer stage, which often guides treatment selection. Systematic evaluation of missing documentation for cancer patients may shed light on where investments can be made to capture more complete data.^17^

In our study, we aim to characterize the impact of unknown documentation across multiple cancer types within a large national cancer registry. Specifically, we examine the prevalence of missing data across the three most common cancer types, and whether characteristics and overall survival of cancer patients with missing data are comparable to those with complete data.

## Methods

We examined the prevalence of patient records with missing data and associated cancer patient overall survival in a large cancer registry commonly used for comparative effective studies in oncology for the three most common cancers in the United States. We compared overall survival differences between patients who have complete versus missing data. This study used de-identified patient information and was granted an exemption by the Yale Human Investigation Committee. This study followed the Strengthening the Reporting of Observational Studies in Epidemiology (STROBE) reporting guidelines.

### Data source

The National Cancer Database (NCDB) is a cancer registry established since the 1980s and jointly sponsored by the American College of Surgeons Commission on Cancer (CoC) and the American Cancer Society.^18^ There are over 130 variables in the NCDB Participant User File (PUF) capturing a range of facility and patient information, tumor characteristics, treatment information, and cancer outcomes that are abstracted by trained tumor registrars.^19,20^ Further details regarding the NCDB are included in the supplement (eMethods).

### Missing data ascertainment

We identified 96 variables which were in use for all diagnosis years and disease sites included in our analysis. From these, we identified variables containing missing data in at least one patient record. Missing data were defined as either “blank” or “unknown” for a variable included in the database. Two clinical oncologists reviewed all variables and excluded variables where blank data entry was allowed by the NCDB data dictionary and may not reflect incomplete clinical documentation. A final 63 variables of interest were identified to compare patients with complete versus missing data (Figure 1, eMethods, eTable 1).

**Table 1.** Percentage of patients with missing data in at least one variable and by variable category.

|  | NSCLC<br>N=1,198,749 | Breast<br>N=2,120,775 | Prostate<br>N=1,158,635 |
| --- | --- | --- | --- |
| Any variable | 71.02% | 54.75% | 39.72% |
| Demographics | 13.01% | 16.25% | 13.94% |
| Cancer Identification | 46.78% | 13.40% | 8.04% |
| Stage | 35.11% | 29.25% | 17.12% |
| Treatment | 16.02% | 19.25% | 12.83% |

**Figure 1.**
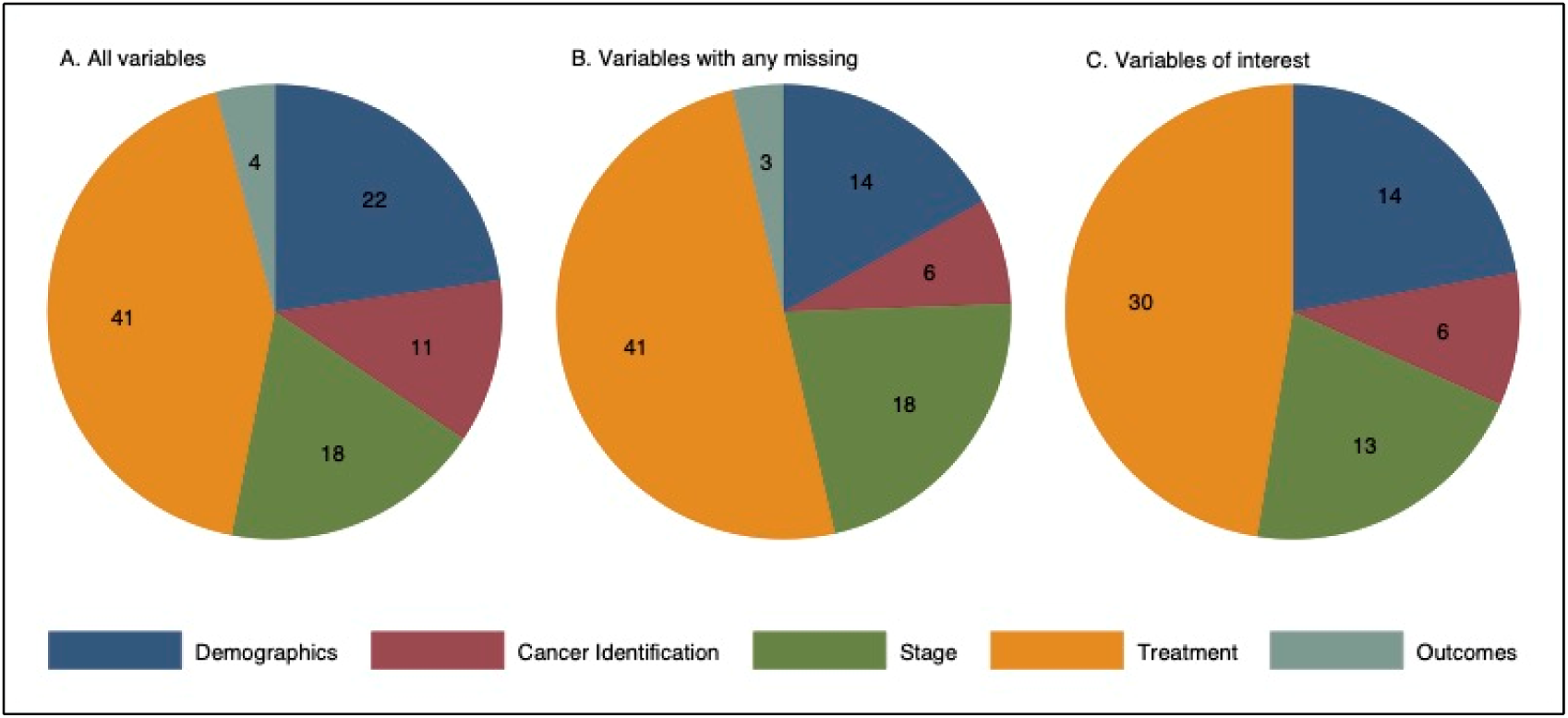
Distribution of variable types among study population.

### Patient selection

We identified non-small cell lung cancer (NSCLC), breast cancer, and prostate cancer patients diagnosed from 2006 to 2015 in the NCDB PUF. Due to changes in data coding rules which introduced new variables and lack of survival information for the most recent diagnosis year, we excluded patients diagnosed in 2016. Given changes in data reporting standards and completeness over time, we examined cancer cases diagnosed in the most recent 10 years prior to 2016. The follow-up period investigated for overall survival included all available follow-up. The impact of missing data was assessed by stage as defined by the NCDB analytic stage group.^21^

### Statistical analysis

We calculated the percentage of patients with missing or unknown values in any 1 of 63 variables of interest. We used standard descriptive statistics, chi-squared test, and Wilcoxon rank-sum test to show differences in patient, tumor, and treatment characteristics between patients with missing versus complete data. Patient records were not used for comparison of patient, tumor, and treatment characteristics if it has a missing value in the variable being compared, which are tabulated in supplemental eTable 2. We used Kaplan-Meier estimates to compare overall survival between patients with missing versus complete data. The primary outcome was the prevalence of missing data and its association with 2-year overall survival. Secondary analysis stratifying by cancer stage and treatment was also performed. Log-rank test was used to identify statistically significant differences in overall survival. We used p<0.05 as the a priori threshold for statistical significance. Hypothesis tests were 2-sided. Bonferroni correction was used to account for multiple comparisons within the study. Threshold for statistical significance for subgroup analysis was p<0.004 after adjustment.

**Table 2.** Patient, disease, and treatment characteristics.

| Non-small cell lung cancer | Patients with<br>complete data<br>N=347,454 | Patients with<br>missing data<br>N=851,295 | p-value |
| --- | --- | --- | --- |
| **Patient and facility** |  |  |  |
| Age at Diagnosis, median (IQR) | 69 (62-76) | 69 (61-77) | <0.001 |
| Sex |  |  | <0.001 |
| Male | 177,594 (51.11%) | 451,217 (53.00%) |  |
| Female | 169,860 (49.89%) | 400,078 (47.00%) |  |
| Race |  |  | <0.001 |
| White | 303,607 (87.38%) | 720,765 (85.59%) |  |
| Black | 34,565 ( 9.95%) | 95,560 (11.35%) |  |
| Other | 9,282 ( 2.67%) | 25,802 ( 3.06%) |  |
| Hispanic Ethnicity |  |  | <0.001 |
| Non-Hispanic | 338,785 (97.50%) | 758,913 (96.80%) |  |
| Hispanic | 8,669 ( 2.50%) | 25,102 ( 3.20%) |  |
| Charlson-Deyo Score |  |  | <0.001 |
| 0 | 184,687 (53.15%) | 503,684 (59.17%) |  |
| 1 | 108,556 (31.24%) | 229,207 (26.92%) |  |
| 2 | 38,916 (11.20%) | 83,537 ( 9.81%) |  |
| >=3 | 15,295 ( 4.40%) | 34,867 ( 4.10%) |  |
| Insurance |  |  | <0.001 |
| Not Insured | 8,818 ( 2.54%) | 27,945 ( 3.38%) |  |
| Private | 92,017 (26.48%) | 226,175 (27.32%) |  |
| Medicaid | 19,886 ( 5.72%) | 53,265 ( 6.43%) |  |
| Medicare | 222,107 (63.92%) | 506,860 (61.22%) |  |
| Other Government | 4,626 ( 1.33%) | 13,691 ( 1.65%) |  |
| Facility Type |  |  | <0.001 |
| Community | 240,682 (69.27%) | 571,663 (67.76%) |  |
| Academic | 106,772 (30.73%) | 271,994 (32.24%) |  |
| **Tumor** |  |  |  |
| Year of Diagnosis, median (IQR) | 2011 (2009-2013) | 2010 (2008-2013) | <0.001 |
| Overall Stage |  |  | <0.001 |
| Stage I | 145,393 (41.85%) | 171,141 (22.04%) |  |
| Stage II | 44,488 (12.81%) | 55,601 ( 7.16%) |  |
| Stage III | 74,441 (21.43%) | 174,460 (22.47%) |  |
| Stage IV | 83,073 (23.91%) | 375,298 (48.33%) |  |
| Tumor Size |  |  | <0.001 |
| <=3 cm | 167,184 (48.19%) | 278,361 (44.46%) |  |
| >3 cm | 179,778 (51.81%) | 347,749 (55.54%) |  |
| Lymph Nodes Involved |  |  | <0.001 |
| No | 197,933 (58.75%) | 287,971 (41.59%) |  |
| Yes | 138,977 (41.25%) | 404,504 (58.41%) |  |
| Distant Metastasis |  |  | <0.001 |
| No | 263,796 (75.92%) | 445,966 (54.69%) |  |
| Yes | 83,658 (24.08%) | 369,411 (45.31%) |  |
| **Treatment** |  |  |  |
| Surgery (Primary Site) |  |  | <0.001 |
| No | 174,754 (50.30%) | 669,039 (78.92%) |  |
| Yes | 172,700 (49.70%) | 178,671 (21.08%) |  |
| Radiation |  |  | <0.001 |
| No | 223,946 (64.45%) | 481,005 (57.20%) |  |
| Yes | 123,508 (35.55%) | 359,919 (42.80%) |  |
| Chemotherapy |  |  | <0.001 |
| No | 207,763 (59.80%) | 434,274 (53.54%) |  |
| Yes | 139,691 (40.20%) | 376,777 (46.46%) |  |
| **Breast cancer** | **Patients with**<br>**complete data** | **Patients with**<br>**missing data** | **p-value** |
|  | N=959,679 | N=1,161,096 |  |
| **Patient and facility** |  |  |  |
| Age at Diagnosis, median (IQR) | 62 (53-72) | 59 (49-70) | <0.001 |
| Sex |  |  | 0.43 |
| Male | 8,552 ( 0.89%) | 10,465 ( 0.90%) |  |
| Female | 951,127 (99.11%) | 1,150,631 (99.10%) |  |
| Race |  |  | <0.001 |
| White | 814,602 (84.88%) | 947,362 (83.24%) |  |
| Black | 105,594 (11.00%) | 137,369 (12.07%) |  |
| Other | 39,483 ( 4.11%) | 53,425 ( 4.69%) |  |
| Hispanic Ethnicity |  |  | <0.001 |
| Non-Hispanic | 915,866 (95.43%) | 982,844 (93.62%) |  |
| Hispanic | 43,813 ( 4.57%) | 66,997 ( 6.38%) |  |
| Charlson-Deyo Score |  |  | <0.001 |
| 0 | 786,312 (81.93%) | 997,133 (85.88%) |  |
| 1 | 137,187 (14.30%) | 131,158 (11.30%) |  |
| 2 | 27,511 ( 2.87%) | 24,880 ( 2.14%) |  |
| >=3 | 8,669 ( 0.90%) | 7,925 ( 0.68%) |  |
| Insurance |  |  | <0.001 |
| Not Insured | 17,384 ( 1.81%) | 25,447 ( 2.27%) |  |
| Private | 486,495 (50.69%) | 626,116 (55.78%) |  |
| Medicaid | 53,951 ( 5.62%) | 70,871 ( 6.31%) |  |
| Medicare | 392,685 (40.92%) | 388,308 (34.59%) |  |
| Other Government | 9,164 ( 0.95%) | 11,747 ( 1.05%) |  |
| Facility Type |  |  | <0.001 |
| Community | 684,570 (71.33%) | 725,684 (67.99%) |  |
| Academic | 275,109 (28.67%) | 341,691 (32.01%) |  |
| **Tumor** |  |  |  |
| Year of Diagnosis, median (IQR) | 2012 (2009-2014) | 2010 (2008-2013) | <0.001 |
| Overall Stage |  |  | <0.001 |
| Stage 0 (DCIS) | 133,409 (13.91%) | 294,752 (27.05%) |  |
| Stage I | 459,031 (47.85%) | 391,027 (35.89%) |  |
| Stage II | 258,213 (26.92%) | 254,836 (23.39%) |  |
| Stage III | 78,254 ( 8.16%) | 97,157 ( 8.92%) |  |
| Stage IV | 30,454 ( 3.17%) | 51,889 ( 4.76%) |  |
| Tumor Size |  |  | <0.001 |
| <=2 cm | 629,447 (65.80%) | 610,410 (63.96%) |  |
| >2 cm | 327,146 (34.20%) | 344,006 (36.04%) |  |
| Lymph Nodes Involved |  |  | <0.001 |
| No | 731,333 (77.35%) | 760,552 (76.73%) |  |
| Yes | 214,178 (22.65%) | 230,644 (23.27%) |  |
| Distant Metastasis |  |  | <0.001 |
| No | 926,319 (96.56%) | 1,030,431 (95.10%) |  |
| Yes | 33,042 ( 3.44%) | 53,149 ( 4.90%) |  |
| **Treatment** |  |  |  |
| Surgery (Primary Site) |  |  | <0.001 |
| No | 46,302 ( 4.82%) | 109,849 ( 9.50%) |  |
| Yes | 913,377 (95.18%) | 1,046,754 (90.50%) |  |
| Radiation |  |  | <0.001 |
| No | 433,200 (45.14%) | 566,736 (49.75%) |  |
| Yes | 526,479 (54.86%) | 572,478 (50.25%) |  |
| Chemotherapy |  |  | <0.001 |
| No | 634,319 (66.10%) | 697,497 (63.81%) |  |
| Yes | 325,360 (33.90%) | 395,557 (36.19%) |  |
| Hormonal Therapy |  |  | <0.001 |
| No | 347,616 (36.22%) | 487,554 (45.72%) |  |
| Yes | 612,063 (63.78%) | 578,864 (54.28%) |  |
| **Prostate cancer** | **Patients with complete data** | **Patient with missing data** | **p-value** |
|  | N=698,468 | N=460,167 |  |
| **Patient and facility** |  |  |  |
| Age at Diagnosis, median (IQR) | 65 (59-71) | 65 (59-72) | <0.001 |
| Sex |  |  |  |
| Male | 698,468 (100.00%) | 460,167 (100.00%) |  |
| Race |  |  | <0.001 |
| White | 579,894 (83.02%) | 361,049 (81.74%) |  |
| Black | 99,417 (14.23%) | 67,160 (15.20%) |  |
| Other | 19,157 ( 2.74%) | 13,501 ( 3.06%) |  |
| Hispanic Ethnicity |  |  | <0.001 |
| Non-Hispanic | 669,071 (95.79%) | 366,527 (94.79%) |  |
| Hispanic | 29,397 ( 4.21%) | 20,141 ( 5.21%) |  |
| Charlson-Deyo Score |  |  | <0.001 |
| 0 | 573,655 (82.13%) | 379,345 (82.44%) |  |
| 1 | 101,891 (14.59%) | 64,092 (13.93%) |  |
| 2 | 17,408 ( 2.49%) | 12,523 ( 2.72%) |  |
| >=3 | 5,514 ( 0.79%) | 4,207 ( 0.91%) |  |
| Insurance |  |  | <0.001 |
| Not Insured | 11,414 ( 1.63%) | 9,344 ( 2.14%) |  |
| Private | 337,278 (48.29%) | 205,477 (47.07%) |  |
| Medicaid | 17,389 ( 2.49%) | 12,835 ( 2.94%) |  |
| Medicare | 318,328 (45.58%) | 201,474 (46.15%) |  |
| Other Government | 14,059 ( 2.01%) | 7,415 ( 1.70%) |  |
| Facility Type |  |  | <0.001 |
| Community | 434,953 (62.27%) | 278,141 (60.56%) |  |
| Academic | 263,515 (37.73%) | 181,155 (39.44%) |  |
| **Tumor** |  |  |  |
| Year of Diagnosis, median (IQR) | 2010 (2008-2013) | 2010 (2007-2012) | <0.001 |
| Overall Stage |  |  | <0.001 |
| Stage I | 96,492 (13.82%) | 48,900 (12.12%) |  |
| Stage II | 493,798 (70.70%) | 266,757 (66.10%) |  |
| Stage III | 73,637 (10.54%) | 43,243 (10.72%) |  |
| Stage IV | 34,503 ( 4.94%) | 44,650 (11.06%) |  |
| Lymph Node Involvement |  |  | <0.001 |
| No | 650,476 (97.24%) | 337,102 (94.65%) |  |
| Yes | 18,464 ( 2.76%) | 19,071 ( 5.35%) |  |
| Distant Metastasis |  |  | <0.001 |
| No | 677,567 (97.01%) | 383,731 (91.83%) |  |
| Yes | 20,862 ( 2.99%) | 34,135 ( 8.17%) |  |
| **Treatment** |  |  |  |
| Surgery (Primary Site) |  |  | <0.001 |
| No | 314,879 (45.08%) | 207,399 (45.39%) |  |
| Yes | 383,589 (54.92%) | 249,492 (54.61%) |  |
| Radiation |  |  | <0.001 |
| No | 446,325 (63.90%) | 303,962 (67.64%) |  |
| Yes | 252,143 (36.10%) | 145,409 (32.36%) |  |
| Chemotherapy |  |  | <0.001 |
| No | 694,105 (99.38%) | 417,776 (98.59%) |  |
| Yes | 4,363 ( 0.62%) | 5,967 ( 1.41%) |  |
| Hormonal Therapy |  |  | <0.001 |
| No | 550,765 (78.85%) | 323,474 (77.11%) |  |
| Yes | 147,703 (21.15%) | 96,039 (22.89%) |  |

Abbreviation: IQR, interquartile range

As a sensitivity analysis, we tested an alternative approach of identifying variables for which data was missing in 1-20% of patient records. This range was determined a priori since <1% missing is likely to have little impact on outcomes of RWD studies, and a large percentage missing is more likely to be reflective of explainable differences in coding rules rather than poor documentation quality. Different percentage thresholds of missing data were also tested (supplemental data). To explore the relative importance of missing data in each individual variable of interest, we also performed univariable Cox regression using a missing indicator for each variable of interest (supplemental data).

Statistical analysis was performed using Stata 16 (StataCorp LLC, College Station, Texas). Our code is available at:https://github.com/Aneja-Lab-Yale/Aneja-Lab-Public-MissingData

## Results

### Distribution of variables and missing data

Of the 96 data elements included for analysis, there were 22 (22.9%) demographics, 11 (11.5%) tumor characteristics, 18 (18.8%) cancer stage, 41 (42.7%) treatment, and 4 (4.2%) outcomes variables. After limiting to variables of interest, there were 14 (22.2%) demographics, 6 (9.5%) tumor characteristics, 13 (20.6%) cancer stage, and 30 (47.6%) treatment variables. (Figure 1). The percentage of patients with missing data in each variable category is shown in Table 1. Differences in patient, tumor, and treatment characteristics between patients with complete and missing data are shown in Table 2.

### Association of missing data with overall survival

For NSCLC, there were 851,295 (71.0%) patients with missing data and 347,454 (29.0%) patients with complete data; 2-year overall survival was 33.2% for patients with missing data and 51.6% for patients with complete data (p<0.001). For breast cancer, there were 1,161,096 (54.7%) patients with missing data and 959,679 (45.3%) patients with complete data; 2-year overall survival was 93.2% for patients with missing data and 93.9% for patients with complete data (p<0.001). For prostate cancer, there were 460,167 (39.7%) patients with missing data and 698,468 (60.3%) patients with complete data; 2-year overall survival was 91.0% for patients with missing data and 95.6% for patients with complete data (p<0.001). This equates to an absolute 2-year overall survival difference of 18.4% for NSCLC, 0.7% for breast cancer, and 4.6% for prostate cancer. (Figure 2)

**Figure 2.**
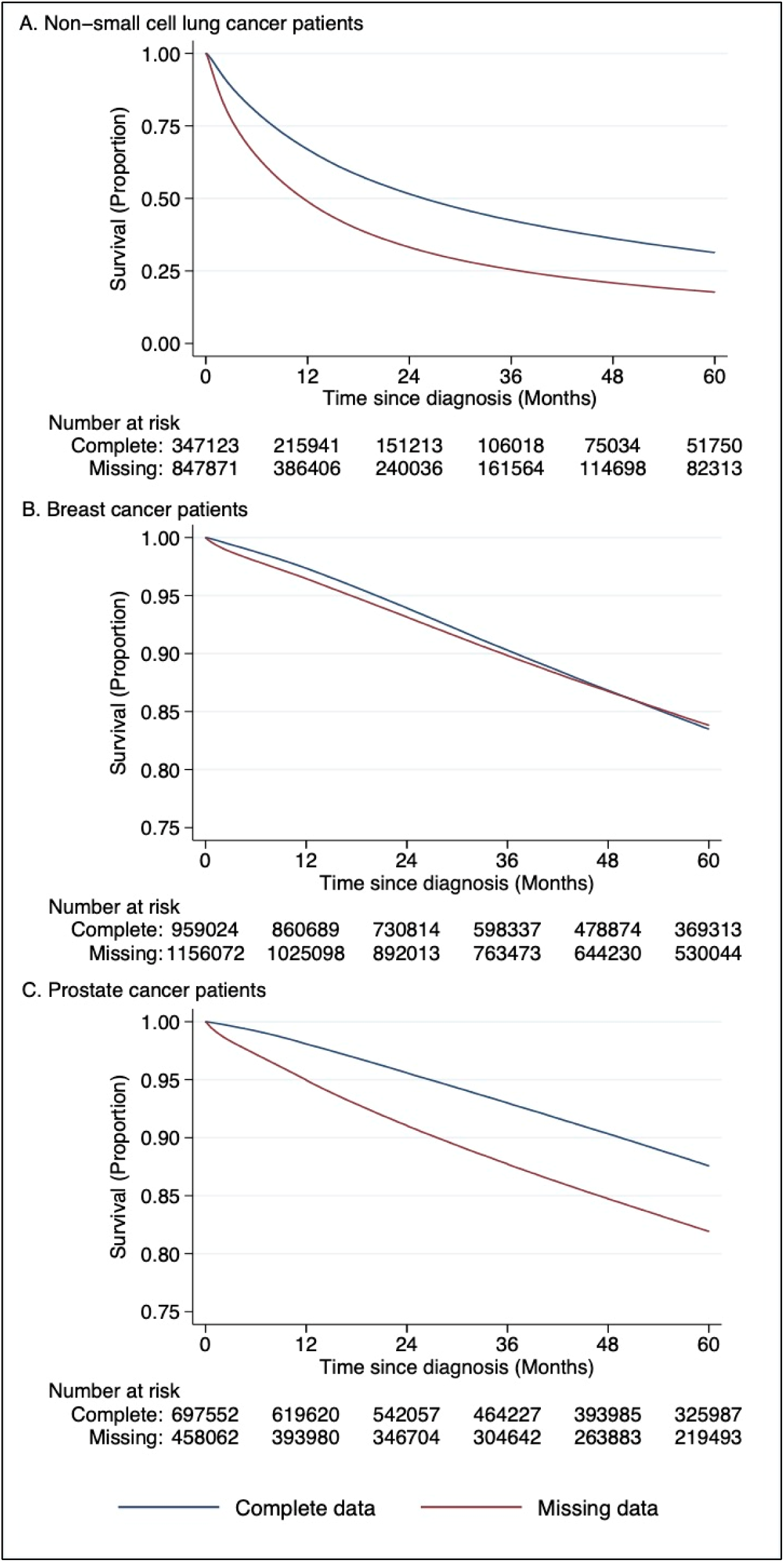
Overall survival of non-small cell lung cancer, breast cancer, and prostate cancer patients by whether data is missing in variables of interest.

Overall survival differences persisted among patients with metastatic disease when stratified by cancer stage. Among non-metastatic patients, the absolute survival differences were smaller for breast (0.4%) and prostate (1.1%) cancer patients, as compared survival differences of 4.5% and 16.7% in metastatic patients respectively (p<0.001 for both, Figure 3). For metastatic NSCLC patients, 2-year overall survival was 13.1% for patients with missing data and 15.0% for patients with complete data (p<0.001); whereas among non-metastatic NSCLC patients, 2-year overall survival was 51.5% for patients with missing data and 63.2% for patients with complete data (p<0.001). Overall survival stratified by cancer stage are shown in eFigures 1-3. Overall survival stratified between receipt of surgery, radiation, or chemotherapy are shown in eFigure 4.

**Figure 3.**
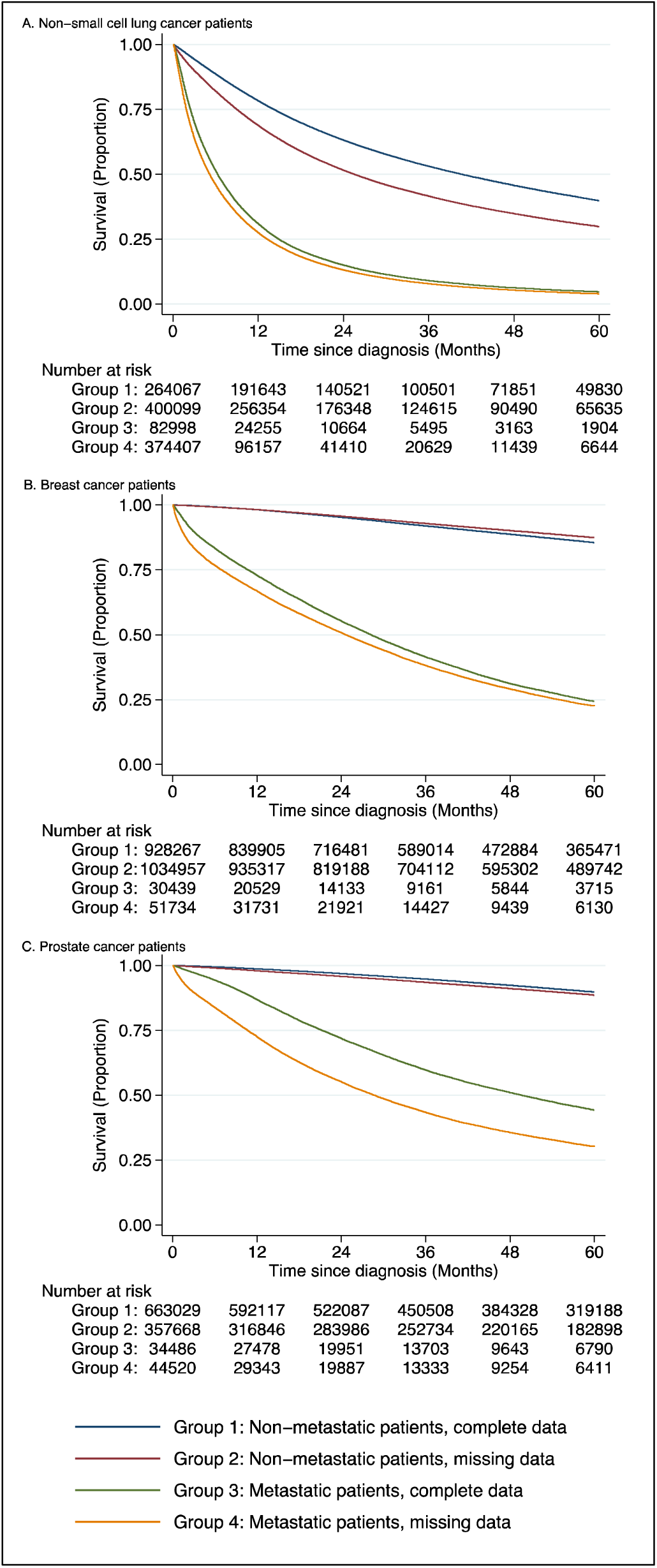
Overall survival of patients with metastatic and non-metastatic non-small cell lung, breast, and prostate cancer by whether data is missing in variables of interest.

### Trends in data completeness and cancer stage over time

There were temporal changes in the proportion of missing data from 2006 to 2015. The percentage of patients with missing data decreased from 81.8% to 67.1% (p<0.001) for NSCLC, from 78.1% to 46.5% (p<0.001) for breast cancer, and from 50.7% to 31.8% (p<0.001) for prostate cancer (eFigure 5). The changes in overall stage are shown in eFigures 6. Overall survival differences stratified by year of diagnosis are shown in eFigure 7.

### Sensitivity analysis using different percentages of missing data

When repeating our analysis using variables for which data was missing in 1-20% of patient records, for NSCLC, there were 622,831 patients with missing data and 575,918 patients without missing data in variables of interest; 2-year overall survival was 33.9% for patients with missing data and 43.5% for patients without missing data (p<0.001). For breast cancer, there were 1,481,729 patients with missing data and 639,046 patients without missing data in variables of interest; 2-year overall survival was 92.4% for patients with missing data and 96.0% for patients without missing data (p<0.001). For prostate cancer, there were 700,523 patients with missing data and 458,112 patients without missing data in variables of interest; 2-year overall survival was 91.7% for patients with missing data and 97.0% for patients without missing data (p<0.001, eFigure 8). Overall survival differences also persisted when we tested different thresholds using either 1-5% or 5-30% missing data as the cutoff (eFigure 9). On exploratory univariable analysis, the relationship between missing data and overall survival differed by individual variables (eTable 3).

## Discussion

In a large national cancer registry, we showed a high prevalence of patient records with missing data in three common cancer types. Missing data was associated with heterogeneous differences in overall survival, and in particular worse overall survival among patients with metastatic disease. The missing data has marked implications for clinical care and research and suggests that there are major gaps in documenting and capturing data via the medical record for patients with cancer.

We showed significant differences in terms of demographics, tumor characteristics, and treatments received between patients with missing data and complete data. Patient records with missing data are more prevalent among blacks and other minorities, reflecting long-standing disparities in access to healthcare and cancer treatment.^22-24^ Patients with fewer comorbid conditions also appeared to more frequently have missing data, which may reflect less available documentation due to fewer medical visits. Advanced stage patients were significantly more likely to have missing data. We hypothesize this is due to increased complexity of care in advanced cancers, leading to increased difficulty in documenting and abstracting all data elements.^25^ The small survival differences in early-stage breast and prostate cancer patients are reflective of this in that definitive and adjuvant therapeutic management options in these settings have relatively less complexity.

Our findings have several implications for clinical care. Missing data is relevant clinically since information that is important for treatment decision making, such as cancer stage, may be incompletely documented. It is also plausible that while a clinical oncologist may have gathered enough information through interviewing and examining the patient, reviewing imaging, consultation with colleagues, or other means, but have not documented this information in the medical record as text that can be abstracted by chart review. In addition, given the multi-disciplinary nature of cancer care, particularly for cases with increased complexity, communication of clinical information between one oncologic specialty to another is often needed to determine the best course of treatment for a patient. However, when a patient’s care is fragmented between institutions, such communication often occurs primarily via sharing of medical records. Therefore, missing data that cannot be abstracted from the medical record have profound implications for cancer patients with fragmented courses of oncologic care. The high prevalence of missing data suggests continued investment into data exchange standards remains a crucial step towards addressing the missing RWD issue for cancer patients.^26,27^

Our findings also have major implications for RWD studies. While incomplete documentation is ubiquitous in RWD sources, observational studies using large cancer registries often exclude patients with missing data, and how missing data is handled is inconsistently reported in the medical literature.^28,29^ Despite an increasing number of papers describing approaches for correcting missing data in observational studies, the practice of handling missing data amongst RWD sources has been slow to change.^30^ Recent systematic comparisons of registry studies and randomized trials do not demonstrate concordant results.^7,9^ Poor quality documentation is therefore a major obstacle to modern RWD sources and can introduce significant biases in research findings using such data, potentially leading to erroneous interpretations regarding real-world clinical outcomes. Within the NCDB, the relative importance of missing data in each individual variable was heterogeneous across cancer types. Variables providing information on staging, diagnosis, and pathology characteristics (such as overall clinical stage, laterality, tumor size and extension, and pathologic lymph node evaluation) appeared to have highly statistically significant associations. Missing values in treatment (such as surgery, radiation) and demographics (such as race, facility type) were also significant. While there are data registry quality control measures, this reflects areas that require ongoing focus to improve the completeness of abstracted data.^31,32^

While generating complete data for all patients is laborious and likely an untenable goal for large cancer registries given the number of patients and variables, there are a number of methods to address missing data within clinical datasets. These include the use of a missing data indicator or simple single value imputation such as replacing missing values with the mean or mode based on non-missing data, which may also introduce bias.^33^ Multiple imputation is an approach less prone to bias compared to single imputation when data is missing at random, but depends on the appropriate modeling of each variable.^34,35^ Recent efforts employing machine learning methods for imputation have shown promise, but often require significant computational resources.^36,37^ There are also ongoing efforts to develop methods for capturing more complete data. For example, greater adherence to structured data entry within the medical record may enable automatic abstraction of structured data elements.^38,39^ For unstructured data, natural language processing tools are being explored to capture information that would otherwise require substantial manual review for data abstraction.^40,41^

Missing data itself may not be the reason for worse survival. The clinical explanations for survival differences associated with missing data are likely multi-factorial. There were significant differences in distribution of cancer stage between patients with and without missing data. The distribution of cancer stage at diagnosis within the NCDB has also changed over time, which has previously been described.^42,43^ Differences in demographic characteristics, year of diagnosis, and treatments received are also contributory factors. There are also likely uncaptured confounders inherent to the observational nature of RWD studies. The decrease in missing data by diagnosis year is reflective of improvements in coding standards and cancer registry quality over time. Our findings are corroborated by other studies examining missing data as a potential source of bias among RWD sources.^44-47^ Our results also corroborate previous analysis showing significant under ascertainment of stage and treatment data within cancer-specific registries.^48-50^ Fragmented care is another plausible explanation for the association between missing data and cancer patient survival.^51^ Since registry data abstraction necessarily depends on information available within the patient record at the reporting facility, documentation quality may particularly affect patients with complex or fragmented disease courses.^52,53^

### Limitations

There are limitations to our analysis. We examined overall survival and cannot draw conclusions on other outcomes such as toxicity, disease recurrence, or causes of death. The dataset within our study is an observational cancer registry, and there may be limitations in the data abstraction process precluding complete capture of the medical record. Patient vital status (alive or dead) is reported to the NCDB from each institution. Given the NCDB does not specify how this is captured at each institution, there may be variability in the capture of overall survival information.^18,32^ However, all RWD sources likely face these limitations to a varying degree, and our analysis therefore should be interpreted as an exemplification of incomplete documentation within RWD sources in oncology. Our study population is also heterogenous. The patients’ cancer treatment paradigms including receipt and sequence of local and systemic therapies necessarily differ and do not reflect one specific clinical scenario. Nevertheless, overall survival differences between patients with missing and complete data persisted despite adjusting for multiple tumor and treatment-related factors. Additionally, the proportion of patients with missing data also depends on the number of variables examined, since it is more difficult to have complete documentation for a larger number of data elements. Given there are a large number of variables within the NCDB, we undertook an alternative analysis of choosing variables with missing data in 1-20% of patient records as variables of interest to identify patients with missing versus complete data. We also tested this assumption in sensitivity analysis, where we show overall survival difference persists using either 1-5% or 5-30% missing as the cutoff.

## Conclusions

In conclusion, we show that the majority of patients in a large cancer registry-based RWD source are subject to missing data. Missing data that was unable to be ascertained from the medical record is associated with heterogenous differences in overall survival, and in particular worse survival among metastatic patients. Increasing documentation quality and adoption of rigorous missing data correction methods are needed to best leverage RWD for clinical advancements.

## Data Availability

The primary data is available by application through the American College of Surgeons (https://www.facs.org/quality-programs/cancer/ncdb/puf). The datasets generated in our analysis can be reproduced using code available at https://github.com/Aneja-Lab-Yale

## Access to Data

Drs. Yang and Aneja had full access to all the data in the study and take responsibility for the integrity of the data and the accuracy of the data analysis.

