## Supplement for "Prevalence of missing data in the National Cancer Database and association with overall survival"

**eMethods**

**eTable 1.** Variables used for all diagnosis years and disease sites included for analysis

**eTable 2.** Count of patient records used for comparison of patient, tumor, and treatment characteristics

**eTable 3.** Univariable Cox regression using a binary indicator variable of whether data is missing in each variable of interest

**eFigure 1.** Non-small cell lung cancer overall survival by whether data is missing in variables of interest and by cancer stage

**eFigure 2.** Breast cancer overall survival by whether data is missing in variables of interest and by cancer stage

**eFigure 3.** Prostate cancer overall survival by whether data is missing in variables of interest and by cancer stage

**eFigure 4.** Overall survival by whether data is missing in variables of interest and by treatment received

**eFigure 5.** Proportion of patients with missing data by year of diagnosis

**eFigure 6.** Distribution of cancer stage by year of diagnosis

**eFigure 7.** Overall survival by whether data is missing in variables of interest and by year of diagnosis

**eFigure 8.** Overall survival by complete versus missing data in variables missing in one to twenty percent of patients

**eFigure 9.** Sensitivity analysis varying percentages of missing data

### eMethods

#### The National Cancer Database

Data reporting to the NCDB follows national registry coding standards and the recording of individual data elements have detailed specifications from the American College of Surgeons Commission on Cancer (CoC). Trained tumor registrars at individual cancer programs abstract specified data elements from patient records in accordance to registry data standards. If a data element is unable to be identified within the patient record, it may be recorded with a blank or unknown value following registry coding guidelines. Specific information abstracted include complex data elements such as comorbidity score, cancer stage, diagnostic procedures, treatment information including receipt of surgery, radiation, chemotherapy, among others.

Data elements in our study were categorized into facility and patient demographics (Demographics), tumor characteristics (Cancer Identification), cancer stage (Stage), cancer treatments (Treatment), and survival outcomes (Outcomes) variables based on the NCDB data dictionary. Two clinical oncologists reviewed all variables and excluded variables where blank data entry was allowed by the NCDB data dictionary and may not reflect incomplete clinical documentation. For example, days from diagnosis to chemotherapy was recorded as missing if the patient never received chemotherapy and therefore excluded from analysis. Furthermore, to be conservative, TNM pathologic staging were not considered missing if pathologic stage was not documented or a pathologic specimen was not collected. A final 63 variables of interest were identified to compare patients with and without completely documented data.

**eTable 1.** Variables used for all diagnosis years and disease sites included for analysis

| Category | Variable name | Missing in at least one patient record | Variable of interest |
|---|---|---|---|
| Demographics | AGE | | |
| Demographics | CDCC_TOTAL_BEST | | |
| Demographics | CROWFLY | X | X |
| Demographics | FACILITY_LOCATION_CD | X | X |
| Demographics | FACILITY_TYPE_CD | X | X |
| Demographics | INSURANCE_STATUS | X | X |
| Demographics | MED_INC_QUAR_00 | X | X |
| Demographics | MED_INC_QUAR_12 | X | X |
| Demographics | MED_INC_QUAR_16 | X | X |
| Demographics | MEDICAID_EXPN_CODE | | |
| Demographics | NO_HSD_QUAR_00 | X | X |
| Demographics | NO_HSD_QUAR_12 | X | X |
| Demographics | NO_HSD_QUAR_16 | X | X |
| Demographics | PUF_CASE_ID | | |
| Demographics | PUF_FACILITY_ID | | |
| Demographics | PUF_MULT_SOURCE | | |
| Demographics | RACE | X | X |
| Demographics | REFERENCE_DATE_FLAG | | |
| Demographics | SEX | | |
| Demographics | SPANISH_HISPANIC_ORIGIN | X | X |
| Demographics | UR_CD_03 | X | X |
| Demographics | UR_CD_13 | X | X |
| Cancer Identification | BEHAVIOR | | |
| Cancer Identification | CLASS_OF_CASE | | |
| Cancer Identification | DIAGNOSTIC_CONFIRMATION | X | X |
| Cancer Identification | GRADE | X | X |
| Cancer Identification | HISTOLOGY | | |
| Cancer Identification | LATERALITY | X | X |
| Cancer Identification | PRIMARY_SITE | | |
| Cancer Identification | REGIONAL_NODES_EXAMINED | X | X |
| Cancer Identification | REGIONAL_NODES_POSITIVE | X | X |
| Cancer Identification | SEQUENCE_NUMBER | X | X |
| Cancer Identification | YEAR_OF_DIAGNOSIS | | |
| Stage | ANALYTIC_STAGE_GROUP | X | X |
| Stage | CS_EXTENSION | X | X |
| Stage | CS_METS_AT_DX | X | X |
| Stage | CS_METS_EVAL | X | X |
| Stage | CS_TUMOR_SIZEEXT_EVAL | X | X |
| Stage | CS_VERSION_LATEST | X | X |

| | | | |
|---|---|---|---|
| Stage | DX_STAGING_PROC_DAYS | X | |
| Stage | RX_SUMM_DXSTG_PROC | X | X |
| Stage | TNM_CLIN_M | X | X |
| Stage | TNM_CLIN_N | X | X |
| Stage | TNM_CLIN_STAGE_GROUP | X | X |
| Stage | TNM_CLIN_T | X | X |
| Stage | TNM_EDITION_NUMBER | X | X |
| Stage | TNM_PATH_M | X | |
| Stage | TNM_PATH_N | X | |
| Stage | TNM_PATH_STAGE_GROUP | X | |
| Stage | TNM_PATH_T | X | |
| Stage | TUMOR_SIZE | X | X[a] |
| Treatment | DX_CHEMO_STARTED_DAYS | X | |
| Treatment | DX_DEFSURG_STARTED_DAYS | X | |
| Treatment | DX_HORMONE_STARTED_DAYS | X | |
| Treatment | DX_IMMUNO_STARTED_DAYS | X | |
| Treatment | DX_OTHER_STARTED_DAYS | X | |
| Treatment | DX_RAD_STARTED_DAYS | X | |
| Treatment | DX_RX_STARTED_DAYS | X | |
| Treatment | DX_SURG_STARTED_DAYS | X | |
| Treatment | DX_SYSTEMIC_STARTED_DAYS | X | |
| Treatment | PALLIATIVE_CARE | X | X |
| Treatment | PALLIATIVE_CARE_HOSP | X | X |
| Treatment | RAD_BOOST_DOSE_CGY | X | X |
| Treatment | RAD_BOOST_RX_MODALITY | X | X |
| Treatment | RAD_ELAPSED_RX_DAYS | X | X |
| Treatment | RAD_LOCATION_OF_RX | X | X |
| Treatment | RAD_NUM_TREAT_VOL | X | X |
| Treatment | RAD_REGIONAL_DOSE_CGY | X | X |
| Treatment | RAD_REGIONAL_RX_MODALITY | X | X |
| Treatment | RAD_TREAT_VOL | X | X |
| Treatment | READM_HOSP_30_DAYS | X | X |
| Treatment | REASON_FOR_NO_RADIATION | X | X |
| Treatment | REASON_FOR_NO_SURGERY | X | X |
| Treatment | RX_HOSP_CHEMO | X | X |
| Treatment | RX_HOSP_DXSTG_PROC | X | X |
| Treatment | RX_HOSP_HORMONE | X | X |
| Treatment | RX_HOSP_IMMUNOTHERAPY | X | X |
| Treatment | RX_HOSP_OTHER | X | X |
| Treatment | RX_HOSP_SURG_PRIM_SITE | X | X |
| Treatment | RX_SUMM_CHEMO | X | X |
| Treatment | RX_SUMM_HORMONE | X | X |
| Treatment | RX_SUMM_IMMUNOTHERAPY | X | X |
| Treatment | RX_SUMM_OTHER | X | X |

| | | | |
|---|---|---|---|
| Treatment | RX_SUMM_RADIATION | X | X |
| Treatment | RX_SUMM_SCOPE_REG_LN_SUR | X | X |
| Treatment | RX_SUMM_SURG_OTH_REGDIS | X | X |
| Treatment | RX_SUMM_SURG_PRIM_SITE | X | X |
| Treatment | RX_SUMM_SURGICAL_MARGINS | X | |
| Treatment | RX_SUMM_SURGRAD_SEQ | X | X |
| Treatment | RX_SUMM_SYSTEMIC_SUR_SEQ | X | X |
| Treatment | RX_SUMM_TRNSPLNT_ENDO | X | X |
| Treatment | SURG_DISCHARGE_DAYS | X | |
| Outcomes | DX_LASTCONTACT_DEATH_MONTHS | X | |
| Outcomes | PUF_30_DAY_MORT_CD | X | |
| Outcomes | PUF_90_DAY_MORT_CD | X | |
| Outcomes | PUF_VITAL_STATUS | | |

[a] Tumor size was not used as a variable of interest for prostate cancer given it is not routinely reported in clinical practice

**eTable 2.** Tabulation of patient records used for comparison of patient, tumor, and treatment characteristics

**A. Non-small cell lung cancer**

| | Records used | | Records not used[a] | |
|---|---|---|---|---|
| | **Complete data** | **Missing data** | **Complete data** | **Missing data** |
| Age at Diagnosis | 347,454 | 851,295 | 0 | 0 |
| Sex | 347,454 | 851,295 | 0 | 0 |
| Race | 347,454 | 842,127 | 0 | 9,168 |
| Hispanic Ethnicity | 347,454 | 784,015 | 0 | 67,280 |
| Charlson-Deyo Score | 347,454 | 851,295 | 0 | 0 |
| Insurance | 347,454 | 827,936 | 0 | 23,359 |
| Facility Type | 347,454 | 843,657 | 0 | 7,638 |
| Year of Diagnosis | 347,454 | 851,295 | 0 | 0 |
| Overall Stage | 347,395 | 776,500 | 59 | 74,795 |
| Tumor Size | 346,962 | 626,110 | 492 | 225,185 |
| Lymph Nodes Involved | 336,910 | 692,475 | 10,544 | 158,820 |
| Distant Metastasis | 347,454 | 815,377 | 0 | 35,918 |
| Surgery (Primary Site) | 347,454 | 847,710 | 0 | 3,585 |
| Radiation | 347,454 | 840,924 | 0 | 10,371 |
| Chemotherapy | 347,454 | 811,051 | 0 | 40,244 |

**B. Breast cancer**

| | Records used | | Records not used | |
|---|---|---|---|---|
| | **Complete data** | **Missing data** | **Complete data** | **Missing data** |
| Age at Diagnosis | 959,679 | 1,161,096 | 0 | 0 |
| Sex | 959,679 | 1,161,096 | 0 | 0 |
| Race | 959,679 | 1,138,156 | 0 | 22,940 |
| Hispanic Ethnicity | 959,679 | 1,049,841 | 0 | 111,255 |
| Charlson-Deyo Score | 959,679 | 1,161,096 | 0 | 0 |
| Insurance | 959,679 | 1,122,489 | 0 | 38,607 |
| Facility Type | 959,679 | 1,067,375 | 0 | 93,721 |
| Year of Diagnosis | 959,679 | 1,161,096 | 0 | 0 |
| Overall Stage | 959,361 | 1,089,661 | 318 | 71,435 |
| Tumor Size | 956,593 | 954,416 | 3,086 | 206,680 |
| Lymph Nodes Involved | 945,511 | 991,196 | 14,168 | 169,900 |
| Distant Metastasis | 959,361 | 1,083,580 | 318 | 77,516 |
| Surgery (Primary Site) | 959,679 | 1,156,603 | 0 | 4,493 |
| Radiation | 959,679 | 1,139,214 | 0 | 21,882 |
| Chemotherapy | 959,679 | 1,093,054 | 0 | 68,042 |
| Hormonal Therapy | 959,679 | 1,066,418 | 0 | 94,678 |

**C. Prostate cancer**

| | Records used | | Records not used | |
|---|---|---|---|---|
| | **Complete data** | **Missing data** | **Complete data** | **Missing data** |
| Age at Diagnosis | 698,468 | 460,167 | 0 | 0 |
| Sex | 698,468 | 460,167 | 0 | 0 |
| Race | 698,468 | 441,710 | 0 | 18,457 |
| Hispanic Ethnicity | 698,468 | 386,668 | 0 | 73,499 |
| Charlson-Deyo Score | 698,468 | 460,167 | 0 | 0 |
| Insurance | 698,468 | 436,545 | 0 | 23,622 |
| Facility Type | 698,468 | 459,296 | 0 | 871 |
| Year of Diagnosis | 698,468 | 460,167 | 0 | 0 |
| Overall Stage | 698,430 | 403,550 | 38 | 56,617 |
| Lymph Nodes Involved | 668,940 | 356,173 | 29,528 | 103,994 |
| Distant Metastasis | 698,429 | 417,866 | 39 | 42,301 |
| Surgery (Primary Site) | 698,468 | 456,891 | 0 | 3,276 |
| Radiation | 698,468 | 449,371 | 0 | 10,796 |
| Chemotherapy | 698,468 | 423,743 | 0 | 36,424 |
| Hormonal Therapy | 698,468 | 419,513 | 0 | 40,654 |

[a] Patient records not used that have complete data are due to recoding of the variable for comparison.

**eTable 3.** Univariable Cox regression for using a binary indicator variable of whether data is missing in each variable of interest

**A. Non-small cell lung cancer**

| Variable | Coefficient | P-Value |
|---|---|---|
| TUMOR_SIZE | 0.75 | <0.001 |
| LATERALITY | 0.70 | <0.001 |
| CS_EXTENSION | 0.64 | <0.001 |
| CS_METS_AT_DX | 0.61 | <0.001 |
| RX_SUMM_SURGRAD_SEQ | 0.58 | <0.001 |
| PALLIATIVE_CARE_HOSP | 0.55 | <0.001 |
| GRADE | 0.54 | <0.001 |
| RAD_LOCATION_OF_RX | 0.51 | <0.001 |
| RAD_TREAT_VOL | 0.48 | <0.001 |
| RAD_REGIONAL_RX_MODALITY | 0.48 | <0.001 |
| REASON_FOR_NO_RADIATION | 0.44 | <0.001 |
| RAD_BOOST_RX_MODALITY | 0.39 | <0.001 |
| RAD_BOOST_DOSE_CGY | 0.38 | <0.001 |
| CS_TUMOR_SIZEEXT_EVAL | 0.33 | <0.001 |
| RAD_ELAPSED_RX_DAYS | 0.33 | <0.001 |
| RAD_REGIONAL_DOSE_CGY | 0.31 | <0.001 |
| REGIONAL_NODES_POSITIVE | 0.31 | <0.001 |
| RAD_NUM_TREAT_VOL | 0.30 | <0.001 |
| TNM_CLIN_STAGE_GROUP | -0.40 | <0.001 |
| FACILITY_TYPE_CD | -0.61 | <0.001 |
| FACILITY_LOCATION_CD | -0.61 | <0.001 |
| TNM_EDITION_NUMBER | -1.00 | <0.001 |
| REGIONAL_NODES_EXAMINED | -0.18 | <0.001 |
| REASON_FOR_NO_SURGERY | 0.26 | <0.001 |
| RX_HOSP_IMMUNOTHERAPY | -0.51 | <0.001 |
| RX_SUMM_IMMUNOTHERAPY | -0.50 | <0.001 |
| RX_HOSP_HORMONE | -0.49 | <0.001 |
| RX_HOSP_CHEMO | -0.33 | <0.001 |
| TNM_CLIN_M | -0.20 | <0.001 |
| RX_SUMM_SYSTEMIC_SUR_SEQ | -0.24 | <0.001 |
| RX_SUMM_RADIATION | -0.33 | <0.001 |
| RX_SUMM_CHEMO | -0.15 | <0.001 |
| RX_SUMM_HORMONE | -0.17 | <0.001 |
| RX_SUMM_TRNSPLNT_ENDO | -0.28 | <0.001 |
| DIAGNOSTIC_CONFIRMATION | 0.44 | <0.001 |
| RX_SUMM_SURG_OTH_REGDIS | -0.29 | <0.001 |
| CS_METS_EVAL | 0.13 | <0.001 |
| SPANISH_HISPANIC_ORIGIN | 0.08 | <0.001 |
| ANALYTIC_STAGE_GROUP | 0.07 | <0.001 |
| PALLIATIVE_CARE | -0.16 | <0.001 |
| READM_HOSP_30_DAYS | -0.07 | <0.001 |
| RX_SUMM_DXSTG_PROC | -0.10 | <0.001 |

| | | |
|---|---|---|
| NO_HSD_QUAR_00 | -0.05 | <0.001 |
| MED_INC_QUAR_00 | -0.05 | <0.001 |
| UR_CD_13 | -0.04 | <0.001 |
| UR_CD_03 | -0.04 | <0.001 |
| RX_HOSP_SURG_PRIM_SITE | 0.17 | <0.001 |
| INSURANCE_STATUS | 0.03 | <0.001 |
| RX_SUMM_SCOPE_REG_LN_SUR | -0.04 | <0.001 |
| RX_SUMM_SURG_PRIM_SITE | -0.07 | <0.001 |
| MED_INC_QUAR_16 | 0.03 | 0.002 |
| RACE | -0.03 | 0.026 |
| NO_HSD_QUAR_16 | 0.02 | 0.082 |
| TNM_CLIN_N | -0.01 | 0.135 |
| TNM_CLIN_T | -0.01 | 0.263 |
| MED_INC_QUAR_12 | 0.02 | 0.295 |
| RX_HOSP_DXSTG_PROC | 0.01 | 0.396 |
| SEQUENCE_NUMBER | 0.04 | 0.683 |
| NO_HSD_QUAR_12 | 0.01 | 0.748 |
| RX_HOSP_OTHER | -0.02 | 0.857 |
| RX_SUMM_OTHER | -0.01 | 0.870 |
| CROWFLY | 0.00 | 0.889 |
| CS_VERSION_LATEST | -22.00 | 0.999 |

**B. Breast cancer**

| Variable | Coefficient | P-Value |
|---|---|---|
| LATERALITY | 1.91 | <0.001 |
| CS_EXTENSION | 1.61 | <0.001 |
| REASON_FOR_NO_SURGERY | 0.91 | <0.001 |
| CS_METS_AT_DX | 0.86 | <0.001 |
| REGIONAL_NODES_POSITIVE | 0.79 | <0.001 |
| CS_TUMOR_SIZEEXT_EVAL | 0.73 | <0.001 |
| RX_SUMM_SURGRAD_SEQ | 0.58 | <0.001 |
| CS_METS_EVAL | 0.54 | <0.001 |
| REGIONAL_NODES_EXAMINED | 0.50 | <0.001 |
| RAD_LOCATION_OF_RX | 0.46 | <0.001 |
| RAD_TREAT_VOL | 0.43 | <0.001 |
| REASON_FOR_NO_RADIATION | 0.37 | <0.001 |
| ANALYTIC_STAGE_GROUP | 0.37 | <0.001 |
| RAD_REGIONAL_RX_MODALITY | 0.41 | <0.001 |
| GRADE | 0.17 | <0.001 |
| FACILITY_TYPE_CD | -0.31 | <0.001 |
| FACILITY_LOCATION_CD | -0.31 | <0.001 |
| TNM_CLIN_STAGE_GROUP | -0.13 | <0.001 |
| RX_SUMM_SYSTEMIC_SUR_SEQ | -0.29 | <0.001 |
| DIAGNOSTIC_CONFIRMATION | 1.42 | <0.001 |
| PALLIATIVE_CARE_HOSP | 0.44 | <0.001 |
| RX_SUMM_IMMUNOTHERAPY | -0.30 | <0.001 |
| RX_HOSP_IMMUNOTHERAPY | -0.31 | <0.001 |
| RX_HOSP_HORMONE | -0.23 | <0.001 |

| | | |
|---|---|---|
| RX_SUMM_HORMONE | -0.16 | <0.001 |
| TNM_EDITION_NUMBER | 0.62 | <0.001 |
| RX_HOSP_SURG_PRIM_SITE | 0.96 | <0.001 |
| RX_SUMM_TRNSPLNT_ENDO | -0.25 | <0.001 |
| RAD_NUM_TREAT_VOL | -0.12 | <0.001 |
| RX_SUMM_SURG_PRIM_SITE | 0.56 | <0.001 |
| RACE | -0.30 | <0.001 |
| RAD_BOOST_RX_MODALITY | 0.11 | <0.001 |
| TNM_CLIN_T | -0.17 | <0.001 |
| RX_SUMM_CHEMO | -0.11 | <0.001 |
| RX_HOSP_CHEMO | -0.15 | <0.001 |
| RX_SUMM_DXSTG_PROC | -0.21 | <0.001 |
| NO_HSD_QUAR_00 | -0.09 | <0.001 |
| MED_INC_QUAR_00 | -0.09 | <0.001 |
| RX_SUMM_SCOPE_REG_LN_SUR | 0.17 | <0.001 |
| READM_HOSP_30_DAYS | -0.08 | <0.001 |
| UR_CD_13 | -0.07 | <0.001 |
| UR_CD_03 | -0.07 | <0.001 |
| RAD_BOOST_DOSE_CGY | 0.06 | <0.001 |
| INSURANCE_STATUS | 0.07 | <0.001 |
| TNM_CLIN_N | 0.09 | <0.001 |
| RX_HOSP_DXSTG_PROC | 0.15 | <0.001 |
| RAD_ELAPSED_RX_DAYS | 0.03 | <0.001 |
| RAD_REGIONAL_DOSE_CGY | -0.03 | <0.001 |
| PALLIATIVE_CARE | 0.07 | 0.009 |
| TUMOR_SIZE | 0.01 | 0.013 |
| MED_INC_QUAR_16 | 0.03 | 0.026 |
| CS_VERSION_LATEST | 1.92 | 0.055 |
| SPANISH_HISPANIC_ORIGIN | 0.01 | 0.098 |
| NO_HSD_QUAR_16 | 0.02 | 0.121 |
| MED_INC_QUAR_12 | 0.04 | 0.169 |
| RX_SUMM_SURG_OTH_REGDIS | 0.03 | 0.173 |
| RX_HOSP_OTHER | 0.15 | 0.262 |
| SEQUENCE_NUMBER | 0.26 | 0.263 |
| RX_SUMM_OTHER | 0.13 | 0.312 |
| RX_SUMM_RADIATION | 0.01 | 0.705 |
| NO_HSD_QUAR_12 | 0.01 | 0.782 |
| TNM_CLIN_M | 0.00 | 0.898 |
| CROWFLY | 0.00 | 0.978 |

**C. Prostate cancer**

| Variable | Coefficient | P-Value |
|---|---|---|
| GRADE | 1.40 | <0.001 |
| TNM_CLIN_T | 0.76 | <0.001 |
| CS_METS_AT_DX | 0.85 | <0.001 |
| CS_EXTENSION | 1.69 | <0.001 |
| CS_TUMOR_SIZEEXT_EVAL | 0.92 | <0.001 |
| REGIONAL_NODES_POSITIVE | 0.57 | <0.001 |

| | | |
|---|---|---|
| ANALYTIC_STAGE_GROUP | 0.30 | <0.001 |
| TNM_CLIN_N | 0.57 | <0.001 |
| RACE | -0.52 | <0.001 |
| TNM_EDITION_NUMBER | 1.07 | <0.001 |
| REASON_FOR_NO_SURGERY | 0.36 | <0.001 |
| RX_HOSP_DXSTG_PROC | 0.72 | <0.001 |
| RX_SUMM_HORMONE | -0.24 | <0.001 |
| REGIONAL_NODES_EXAMINED | 0.23 | <0.001 |
| RX_HOSP_HORMONE | -0.30 | <0.001 |
| CS_METS_EVAL | 0.20 | <0.001 |
| DIAGNOSTIC_CONFIRMATION | 0.59 | <0.001 |
| REASON_FOR_NO_RADIATION | 0.16 | <0.001 |
| RX_SUMM_DXSTG_PROC | 0.29 | <0.001 |
| RX_SUMM_SURG_OTH_REGDIS | -0.30 | <0.001 |
| PALLIATIVE_CARE | -0.33 | <0.001 |
| RX_HOSP_IMMUNOTHERAPY | -0.19 | <0.001 |
| RX_SUMM_IMMUNOTHERAPY | -0.19 | <0.001 |
| RX_SUMM_TRNSPLNT_ENDO | -0.16 | <0.001 |
| SPANISH_HISPANIC_ORIGIN | -0.07 | <0.001 |
| RX_HOSP_SURG_PRIM_SITE | 0.54 | <0.001 |
| TNM_CLIN_STAGE_GROUP | 0.05 | <0.001 |
| NO_HSD_QUAR_00 | -0.10 | <0.001 |
| RAD_REGIONAL_DOSE_CGY | 0.11 | <0.001 |
| MED_INC_QUAR_00 | -0.10 | <0.001 |
| RX_SUMM_CHEMO | -0.10 | <0.001 |
| RX_SUMM_SURGRAD_SEQ | 0.15 | <0.001 |
| RAD_BOOST_RX_MODALITY | 0.11 | <0.001 |
| RAD_BOOST_DOSE_CGY | 0.10 | <0.001 |
| RAD_TREAT_VOL | 0.11 | <0.001 |
| RAD_ELAPSED_RX_DAYS | 0.08 | <0.001 |
| RAD_REGIONAL_RX_MODALITY | 0.10 | <0.001 |
| UR_CD_03 | -0.08 | <0.001 |
| UR_CD_13 | -0.08 | <0.001 |
| RX_SUMM_RADIATION | -0.12 | <0.001 |
| RAD_LOCATION_OF_RX | 0.09 | <0.001 |
| RX_SUMM_SYSTEMIC_SUR_SEQ | -0.06 | <0.001 |
| RAD_NUM_TREAT_VOL | 0.05 | <0.001 |
| TNM_CLIN_M | 0.05 | <0.001 |
| FACILITY_TYPE_CD | -0.33 | 0.001 |
| FACILITY_LOCATION_CD | -0.33 | 0.001 |
| RX_SUMM_SCOPE_REG_LN_SUR | -0.07 | 0.008 |
| READM_HOSP_30_DAYS | 0.04 | 0.027 |
| MED_INC_QUAR_12 | 0.07 | 0.064 |
| NO_HSD_QUAR_12 | 0.07 | 0.098 |
| RX_HOSP_CHEMO | -0.03 | 0.099 |
| INSURANCE_STATUS | 0.02 | 0.123 |
| RX_SUMM_SURG_PRIM_SITE | 0.06 | 0.161 |

| | | |
|---|---|---|
| RX_SUMM_OTHER | 0.22 | 0.217 |
| RX_HOSP_OTHER | 0.22 | 0.228 |
| PALLIATIVE_CARE_HOSP | -0.04 | 0.235 |
| SEQUENCE_NUMBER | 0.34 | 0.241 |
| CROWFLY | 0.06 | 0.244 |
| NO_HSD_QUAR_16 | -0.01 | 0.673 |
| MED_INC_QUAR_16 | 0.00 | 0.821 |
| CS_VERSION_LATEST | -21.00 | 0.999 |
| LATERALITY[a] | - | - |

[a] There were no missing values for laterality for prostate cancer. Prostate is not a paired site.

**eFigure 1.** Non-small cell lung cancer overall survival by whether data is missing in variables of interest and by cancer stage

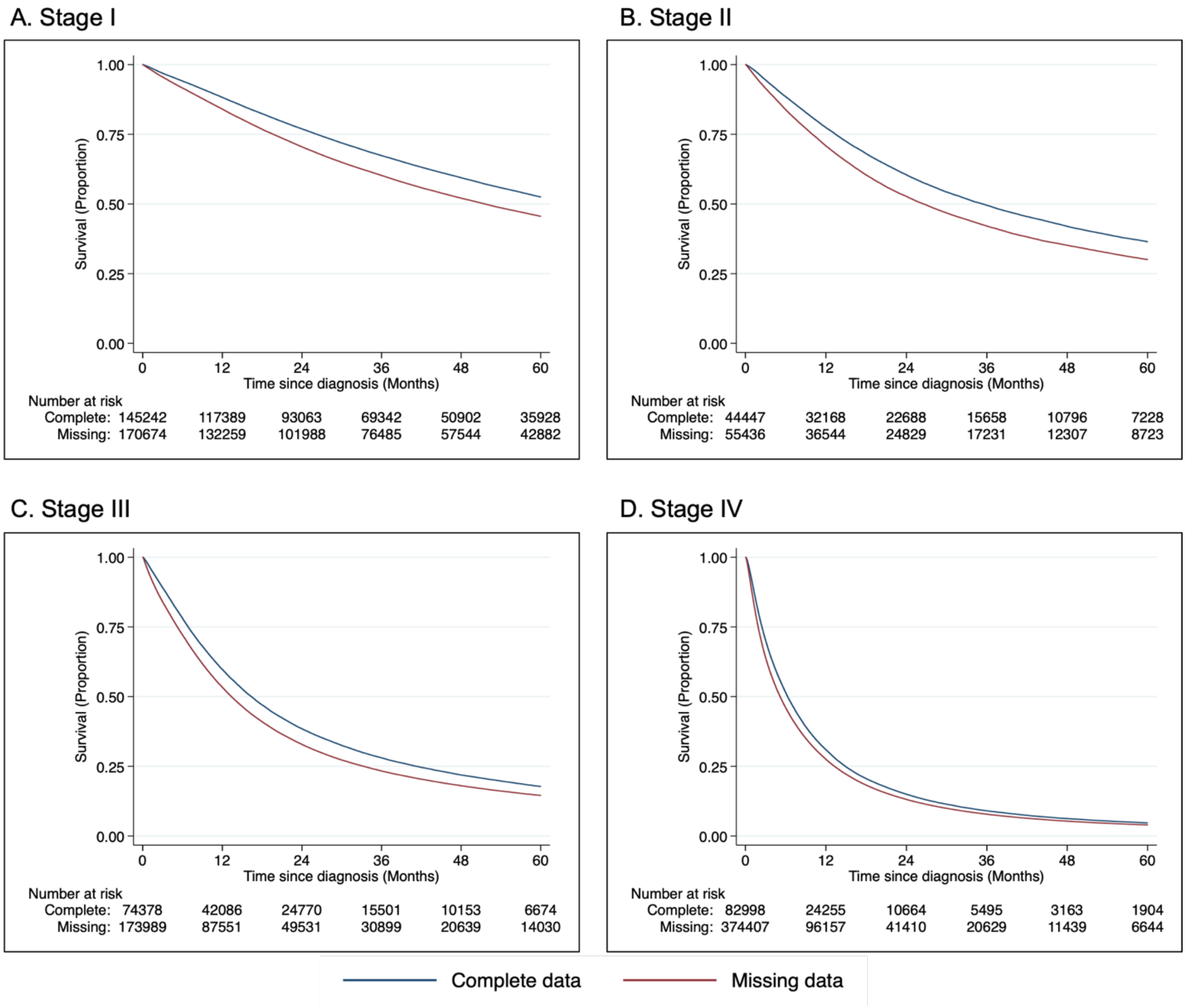


Analytic group stages 0 and I were combined for stage I subgroup analysis for non-small cell lung cancer

**eFigure 2.** Breast cancer overall survival by whether data is missing in variables of interest and by cancer stage

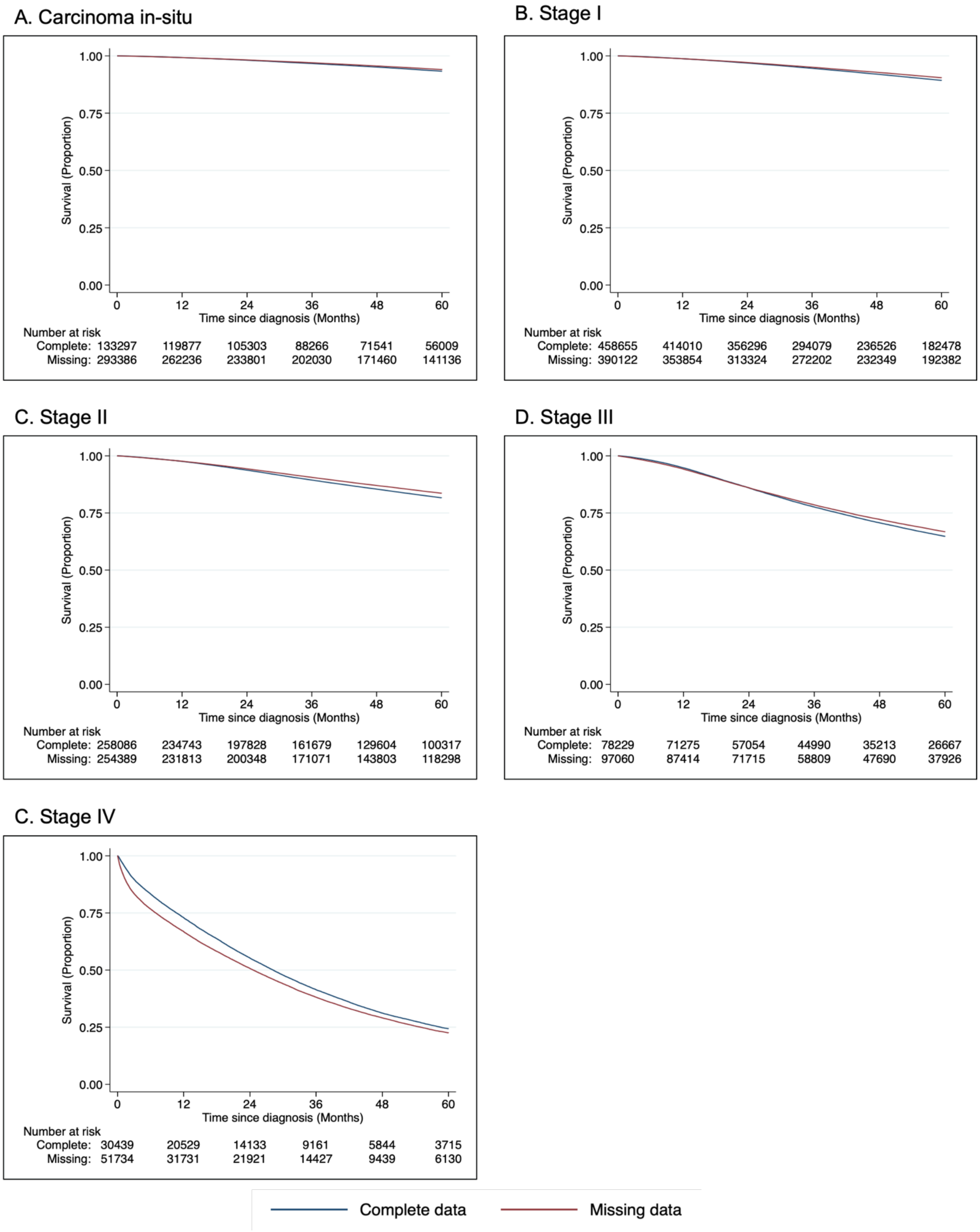

**eFigure 3.** Prostate cancer overall survival by whether data is missing in variables of interest and by cancer stage

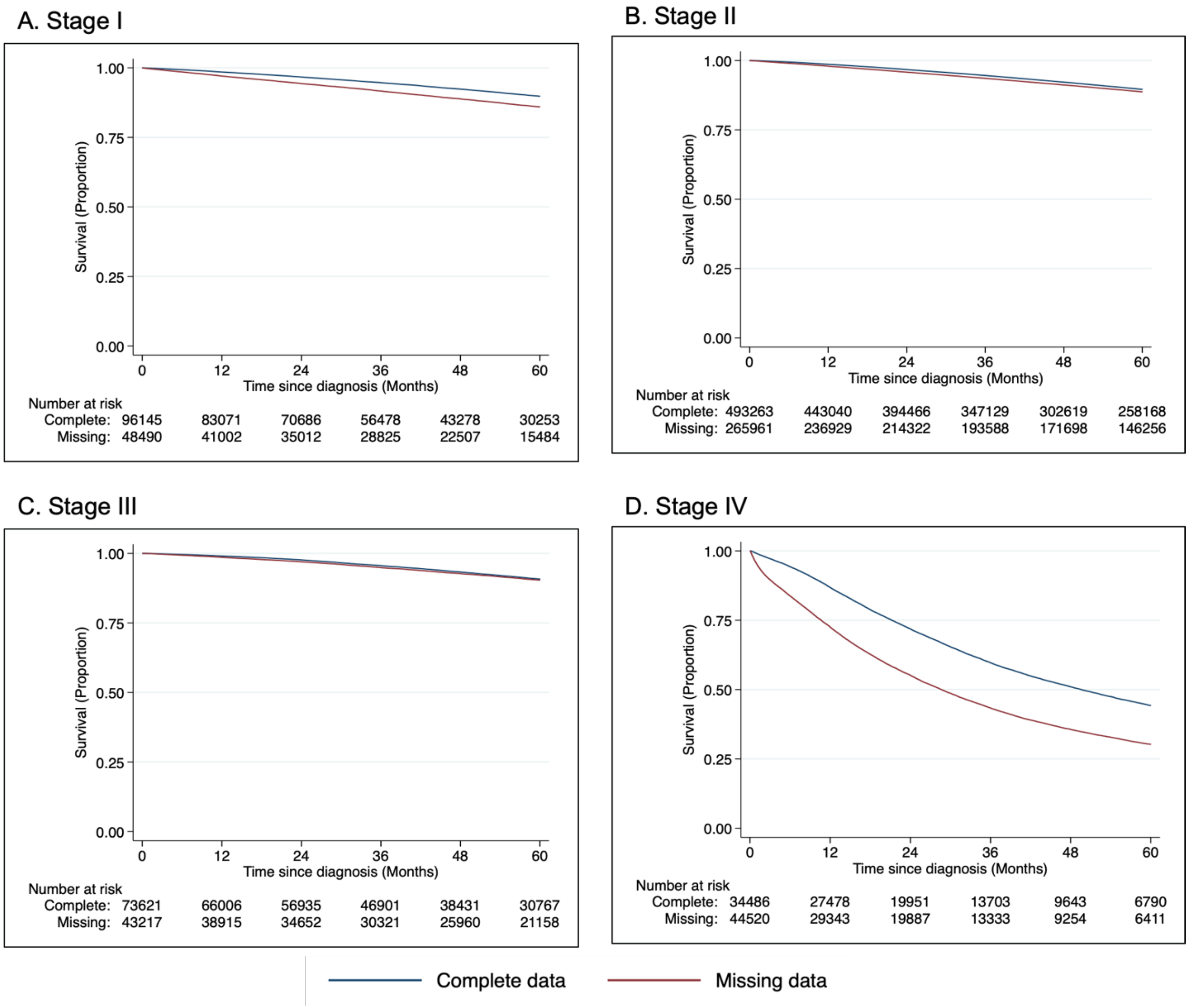

**eFigure 4.** Overall survival by whether data is missing in variables of interest and by treatment received

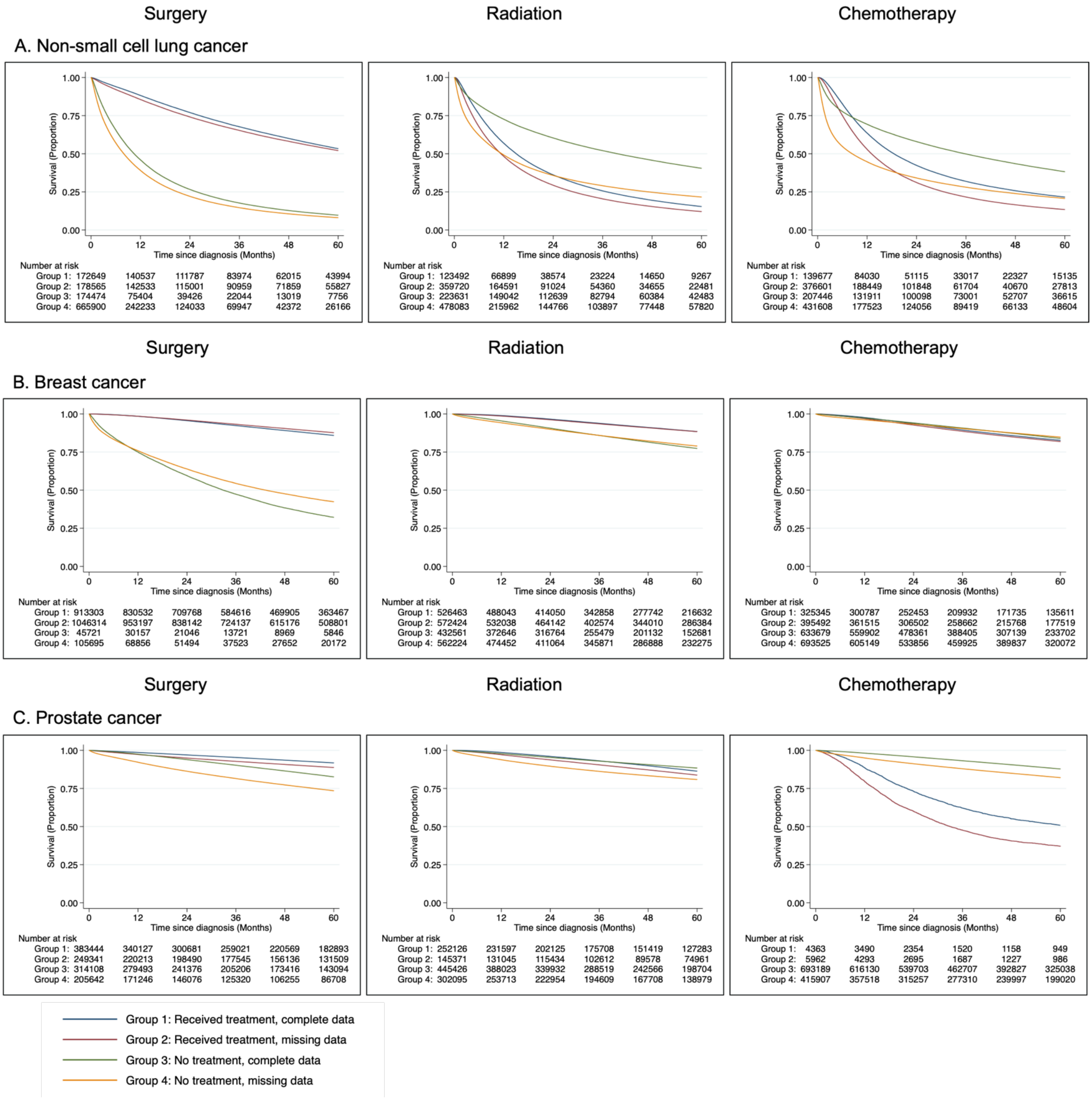

**eFigure 5.** Proportion of patients with missing data by year of diagnosis

A. Non-small cell lung cancer

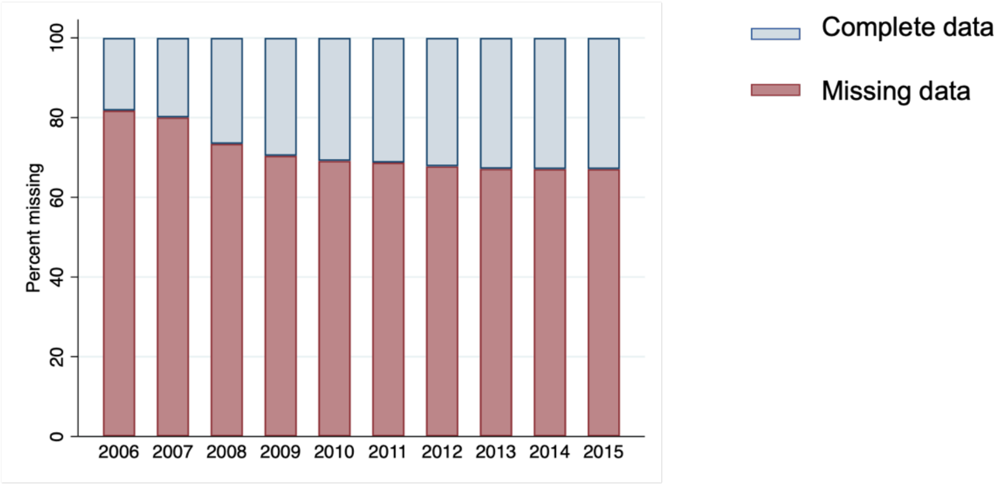


B. Breast cancer

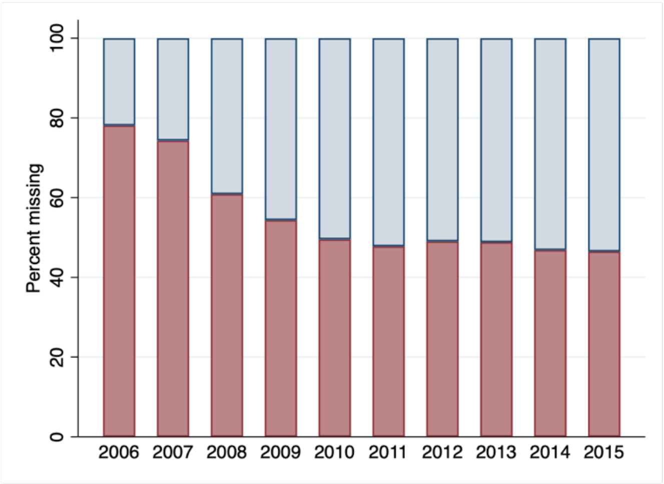


C. Prostate cancer

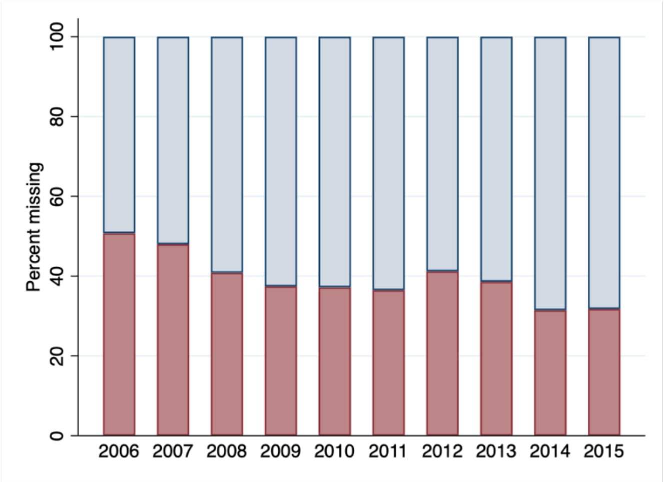

**eFigure 6.** Distribution of cancer stage by year of diagnosis

A. Non-small cell lung cancer

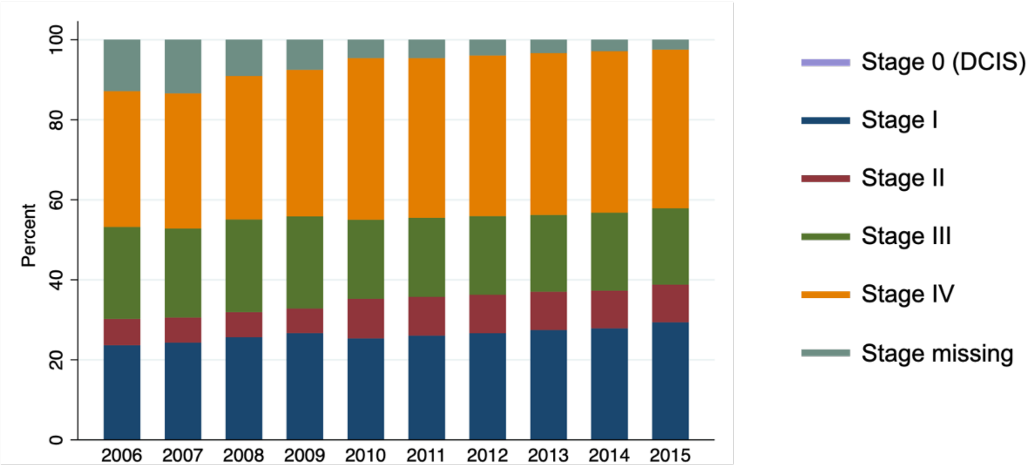


B. Breast cancer

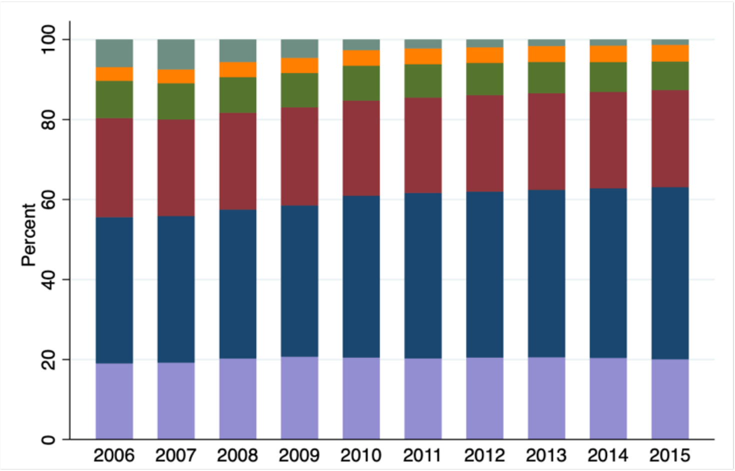


C. Prostate cancer

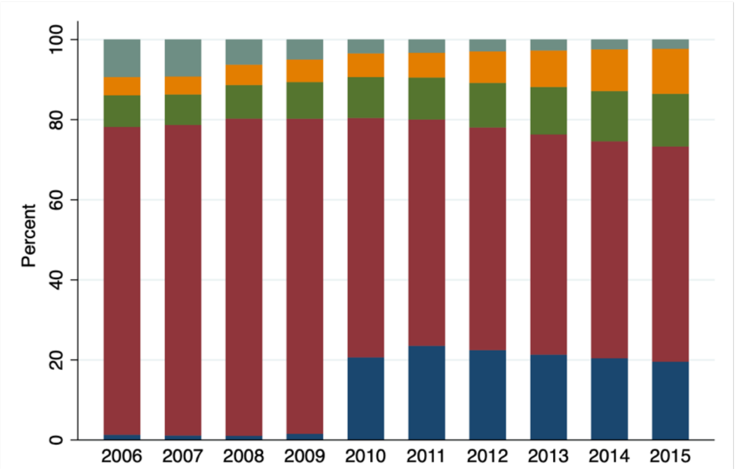

**eFigure 7.** Overall survival by whether data is missing in variables of interest and by year of diagnosis

Diagnosed between 2006 to 2010

Diagnosed between 2011 to 2015

A. Non-small cell lung cancer

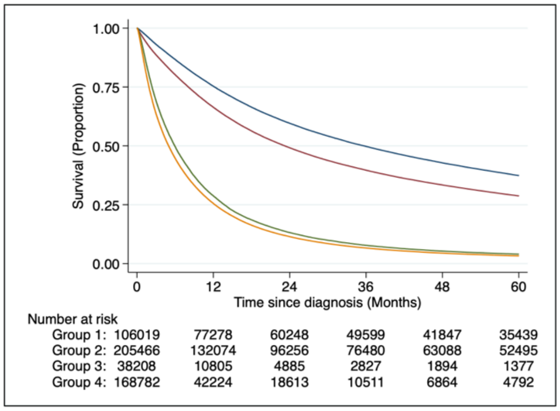


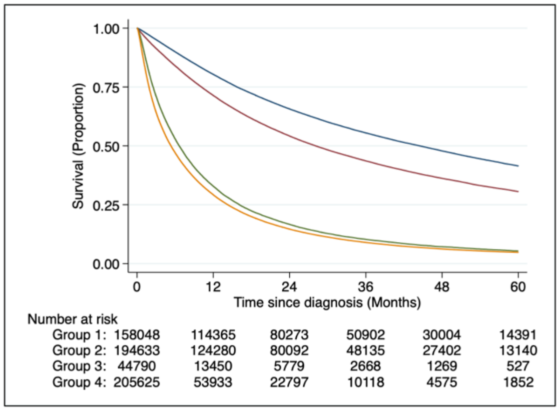


B. Breast cancer

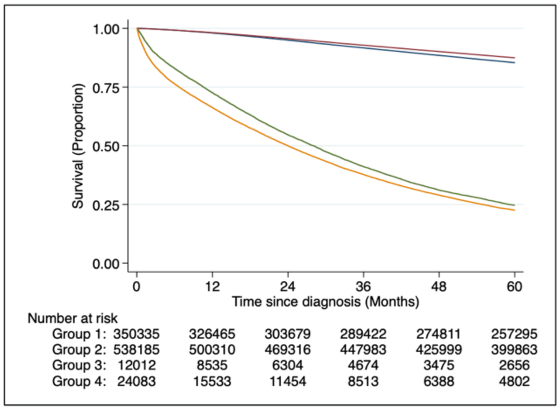


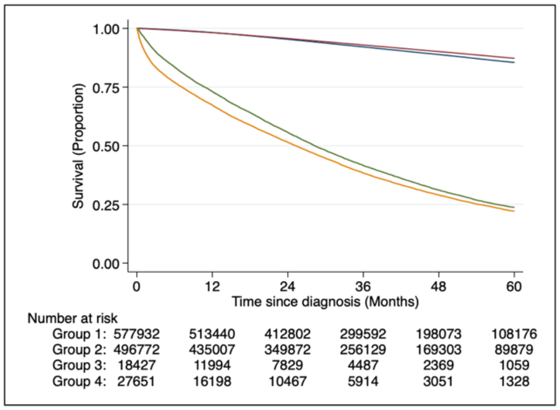


C. Prostate cancer

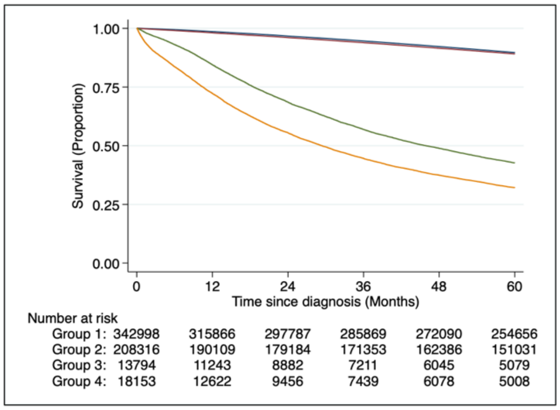


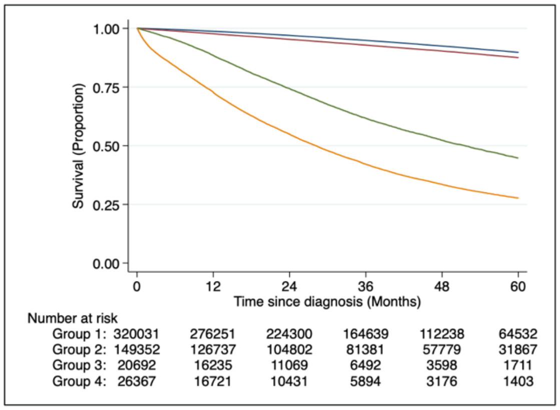


Group 1: Non–metastatic patients, complete data
Group 2: Non–metastatic patients, missing data
Group 3: Metastatic patients, complete data
Group 4: Metastatic patients, missing data

**eFigure 8.** Overall survival by complete versus missing data in variables missing in one to twenty percent of patients

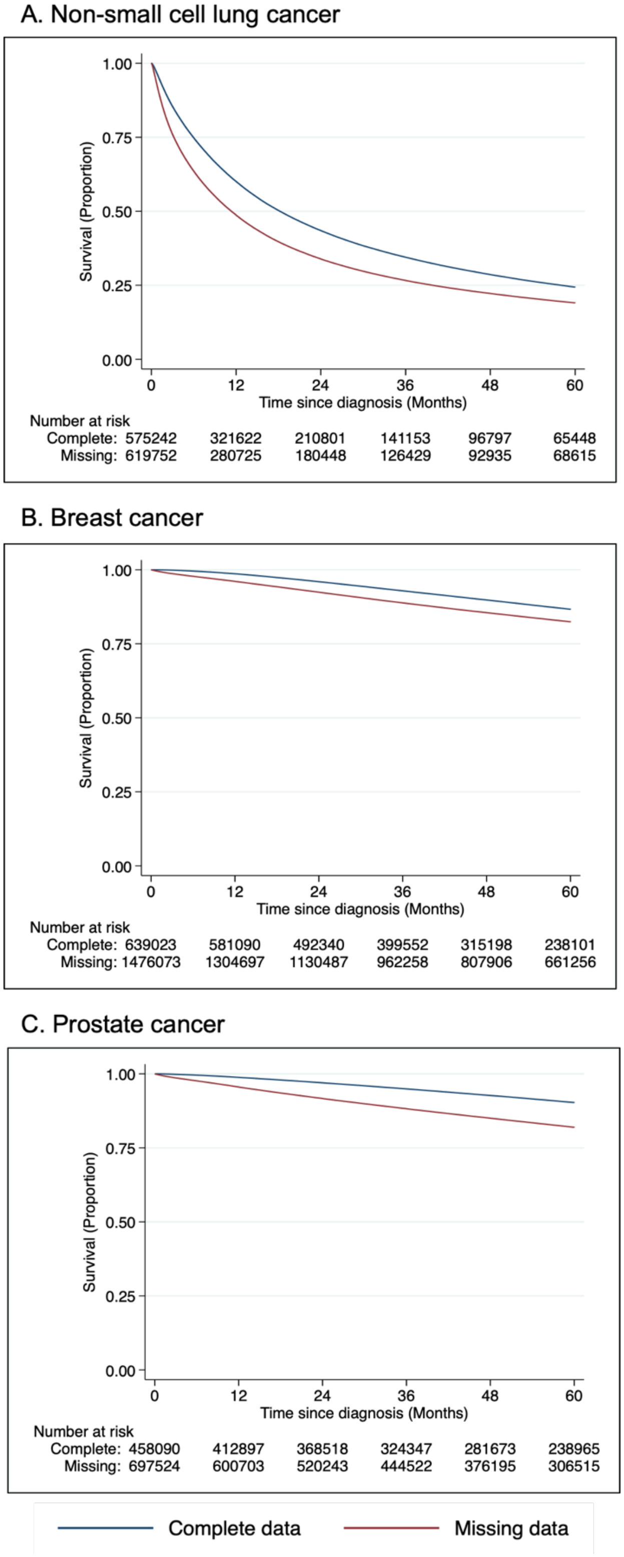

**eFigure 9.** Sensitivity analysis varying percentages of missing data

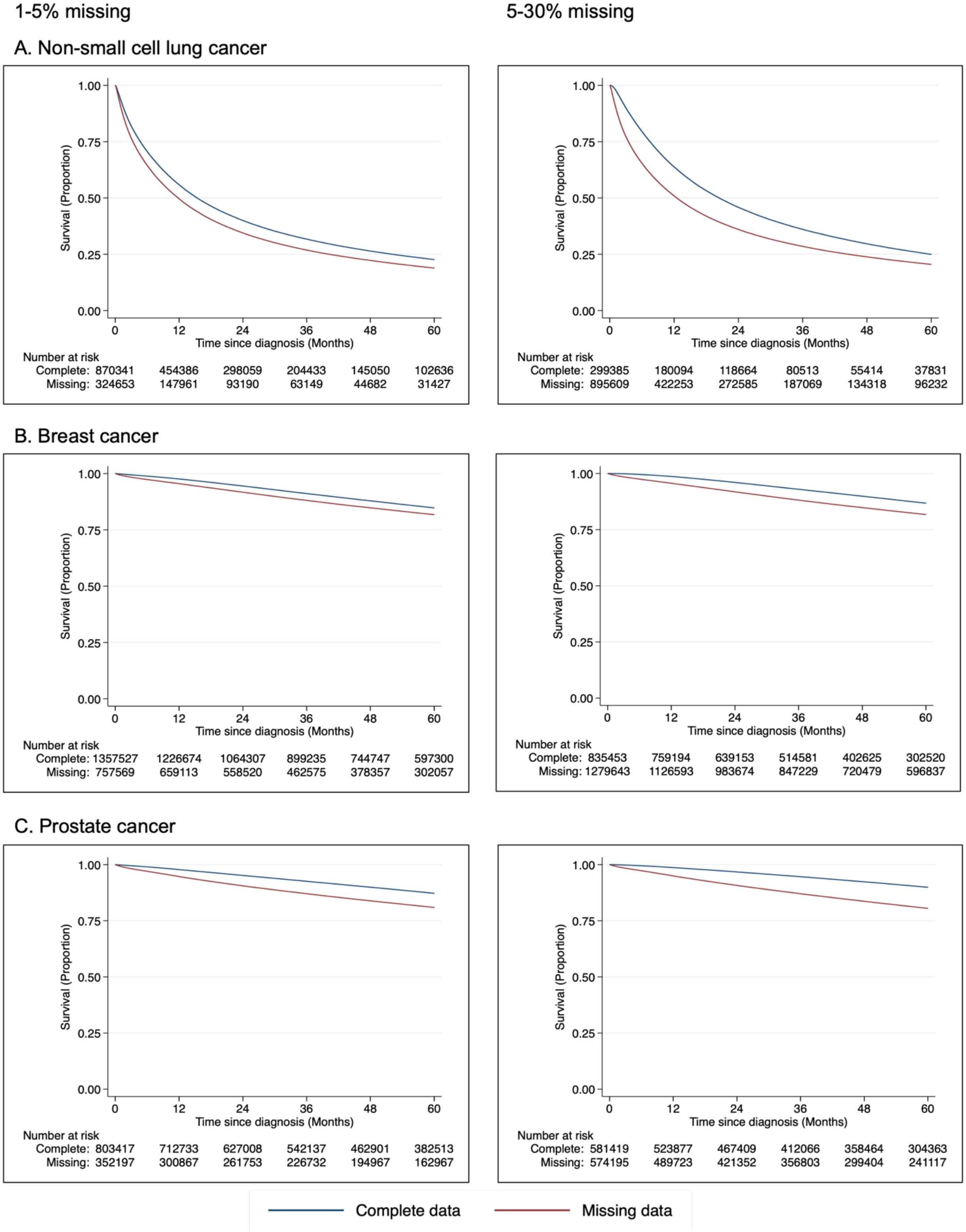
